# Current Knowledge on Potential Determinants of Mass Public Shooting Perpetration and Casualties: A Systematic Review

**DOI:** 10.1101/2022.06.30.22277119

**Authors:** Wit Wichaidit

## Abstract

**Background:** In the United States, mass shootings can be regarded as a public health issue due to its ubiquitous and public nature. A systematic review of the literature with adoption of the social ecological model for interpretation of the evidence can help inform relevant stakeholders and the public regarding the coherence between proposed gun control legislations and the body of empirical evidence.

**Aims:** To conduct a systematic review of factors associated with: 1) perpetration of mass shooting events, and; 2) injuries and fatalities at mass shooting events.

**Material and Methods:** The author conducted a search of the literatures on PubMed using the term “mass shooting”, filtering manuscripts from 2017 to 2022. The author included only articles in peer-reviewed academic journals with primary data collection for the review.

**Results:** The author reviewed 36 of the 336 articles initially found from a search on PubMed. All but two studies focused on mass shootings in the United States. Factors associated with perpetration of mass shootings included having mental illness and experience of acute life stressors (intra-personal), relationship issues with family and friends (inter-personal), gun ownership and access (community), permissive firearm laws or lack of gun control (policy). Factors associated with injuries and fatalities at mass shootings included use of semi-automatic rifles (intrapersonal), targeting women (interpersonal), presence of armed officers at schools (organizational), and lack of gun control measures (policy).

**Conclusion:** The author found that easy access to high-power firearms and lack of gun control were associated with both mass shooting perpetration and casualty. However, caveats including a limited time frame, limited sources of publications, and subjectivity in building the model should be considered in the interpretation of the study findings.

## INTRODUCTION

On 14 May 2022, an 18-year-old male entered Tops Friendly Markets supermarket in the city of Buffalo, New York, USA, and opened fire with a Bushmaster AR-15 style semi-automatic rifle and killed 10 persons, apparently motivated by white supremacy (McKinley et al., 2022). Ten days later, another 18-year-old male entered Robb Elementary School in the city of Uvalde, Texas, USA, and opened fire with a Daniel Defense AR-15 style semi-automatic rifle and killed 19 schoolchildren and 2 teachers (Pilkington, 2022). These incidents are known as mass shootings, and are typically defined as incidents of gun violence not related to armed conflicts or domestic violence that resulted in three or more casualties (injuries or deaths), not including the perpetrator(s). Although mass shootings are not limited to any particular country, the United States is the only country in the OECD where public mass shootings occur every single year, and account for over 70% of all mass shootings in high-income countries (Silva, 2022). Furthermore, mass shootings in the United States tend to happen in public places. For example, the deadliest incident was the Las Vegas shooting in which a 64-year-old male in possession of 24 rifles opened fire on Route 91 Harvest country music festival attendees from his hotel room window and killed 58 people and injured more than 500 persons(Keneally, 2019).

Considering that gun manufacturing and import has risen dramatically in the past decade (Bump 2022) and that civilian firearm ownership is currently the highest in the world at 120 firearms per 100 residents (Karp, 2018), one can argue that there is no population sub-group that is invincible from gun violence and mass shootings. Thus, mass shootings (and, by extension, gun violence) should be regarded as a public health issue in the United States. As a public health issue, mass shooting prevention efforts thus should entail both primary prevention (i.e., prevention of perpetration of mass shootings) and secondary prevention (i.e., minimization of casualties – injuries and deaths – at mass shooting events). Using the lens of public health, perpetration and casualties of mass shootings should be regarded not only as attributable to the perpetrator’s own characteristics, but also to social and environmental factors. Thus, the social ecological model for health behaviors (McLeroy et al., 1988) is potentially useful for reviewing the determinants of mass shootings, which can help relevant stakeholders consider the phenomenon in a more comprehensive manner.

In response to the incidents in Uvalde and the surge in frequency of mass shootings, US legislators have made a bipartisan agreement on federal-level gun control (Williams, 2022). A systematic review on determinants and casualties of mass shootings with interpretation using the social ecological model can potentially help inform relevant stakeholders and the public with regard to the coherence between proposed gun control legislations and the body of empirical evidence, and whether there are additional determinants of mass shootings that should be considered in future agendas. The objective of this study was to conduct a systematic review of factors associated with: 1) perpetration of mass shooting events, and; 2) injuries and fatalities at mass shooting events.

## METHODS

The author conducted a search of the literature on PubMed Central (PMC) on 2 June 2022. Searches were limited to articles published in English from 1 January 2017 to the date of the search. The author used only one search term (“mass shooting”). The author decided to use 2017 as the starting year in an arbitrary manner, as it was the year with the deadliest mass shooting in US history occurred.

The author then excluded duplicates, screened the title of searched articles, excluded articles with no abstract or full text available, and then individually assessed each article for eligibility to review in full. The process and number of publications excluded in each step can be found in *Figure 1*. The author then reviewed the content of the eligible articles and summarized the findings, assigning a unique ID to each article for ease of reference during interpretation and summary.

**Figure 1.**
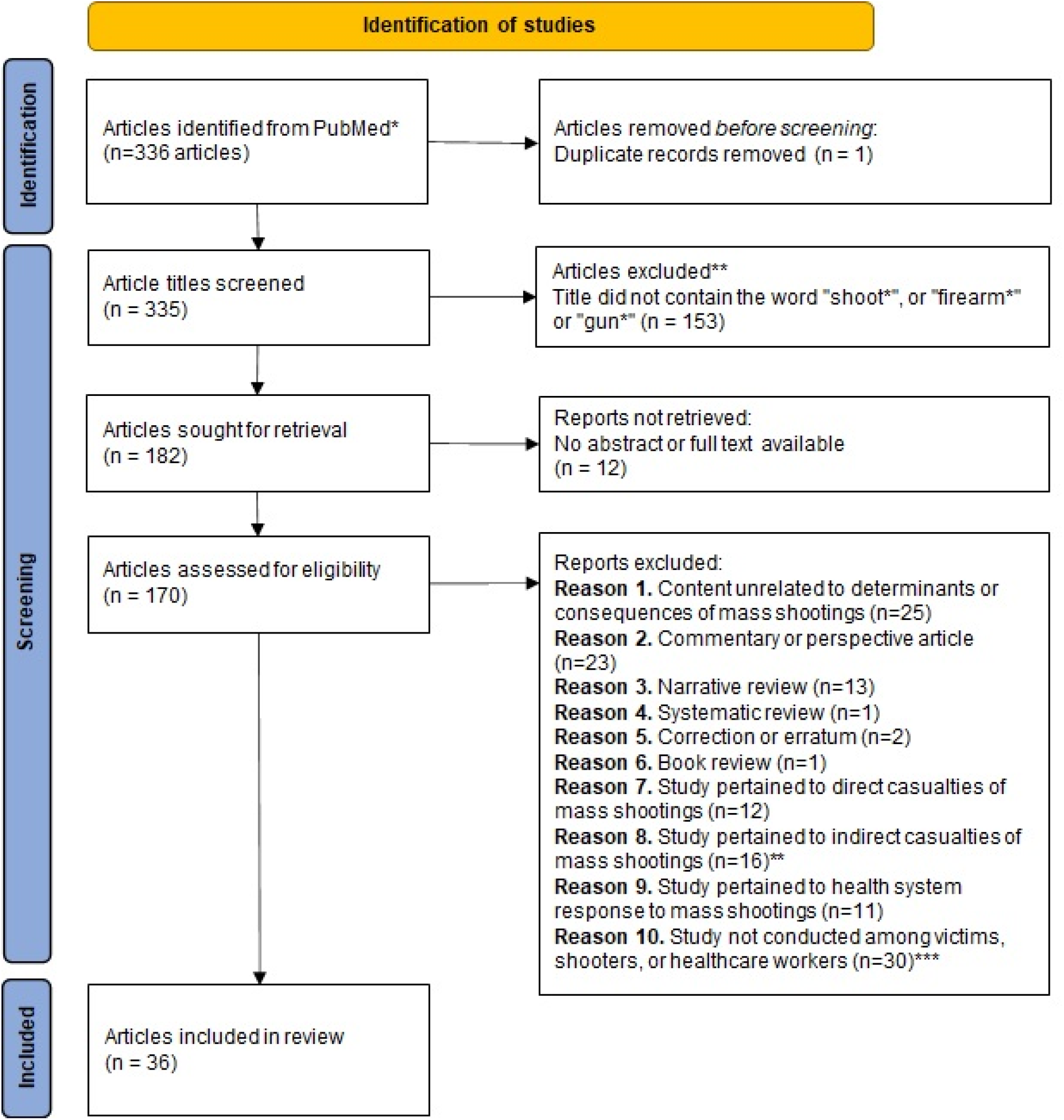
PRISMA 2020 flow diagram for inclusion of articles indexed in PubMed for the review. *Search term “mass shooting”, filter applied to within the last 5 years (2017-2022) **Including family or classmates of direct casualties and individuals affected by vicarious trauma ***Including but not limited to media studies, changes in public opinion, changes in gun ownership

The author then applied the social ecological model (McLeroy et al., 1988) to identify factors associated with perpetration, injuries, and fatalities of mass shooting events, with separate analyses for factors associated with perpetration of mass shooting events, and factors associated with casualties (injuries and fatalities) of mass shooting events. This systematic review did not involve any data collection from human subjects.

## RESULTS

### Study flow and characteristics of included studies

The author initially found 336 articles from a search on PubMed, 1 article was a duplicate and thus removed (Figure 1). The author screened the title of the remaining 335 articles and excluded 153 articles that did not contain the word “shoot*”, “firearm*”, or “gun*” in the title. The author then attempted to retrieve the remaining 182 articles, 12 of which did not have an abstract or full text available. The author assessed the remaining 170 articles for eligibility, and excluded 134 articles for varying reasons, and included 36 research articles in the review and assessment with social ecological model.

All but two of the articles reviewed focused on mass shootings in the United States: one article analyzed non-war mass murders worldwide (including those not involving firearms), another article examined Australian mass shooting incidents and offenders from 1964 to 2014. All articles were ecological, cross-sectional, or panel studies. One study that described itself as a “cohort study” did not follow a defined cohort, but rather examined the location of mass shootings relative to schools and places frequented by children.

### Factors associated with perpetration of mass shooting

Intrapersonal factors associated with mass shootings included having mental illness and experience of acute life stressor (*Table 2*). A number of psychological and behavioral issues seemed to also be common among mass shootings, including interest in past mass killings, self-estrangement, online radicalization, and gun ownership. At the interpersonal level, perpetrators of mass shooters tended to have relationship issues and experienced estrangement from family, friends, and classmates. Surprisingly, a study found that mass shooters were less likely to be single and more likely to have children or be separated or divorced. Household-level gun ownership was also positively associated with perpetration of mass shootings.

**Table 1.**
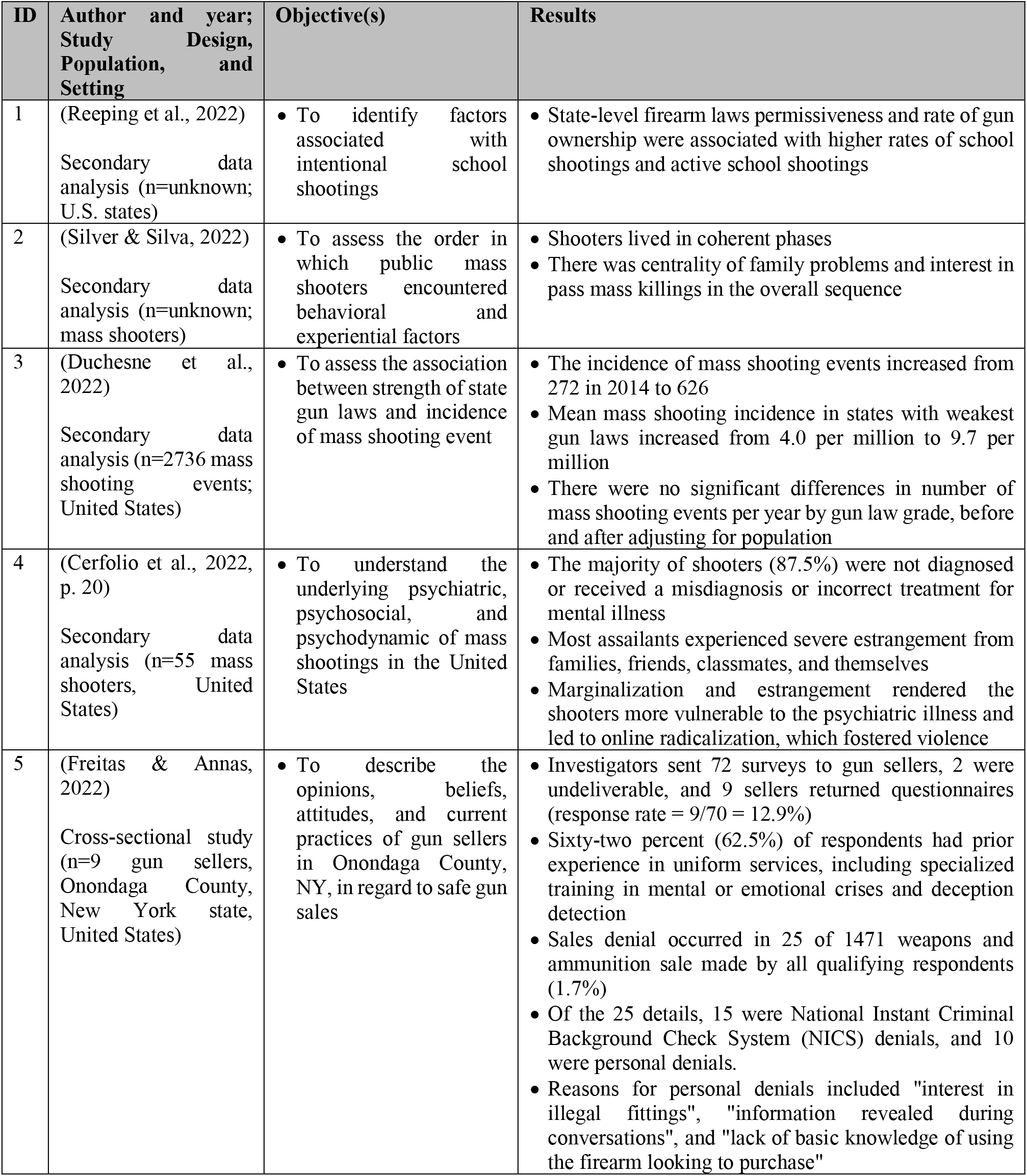
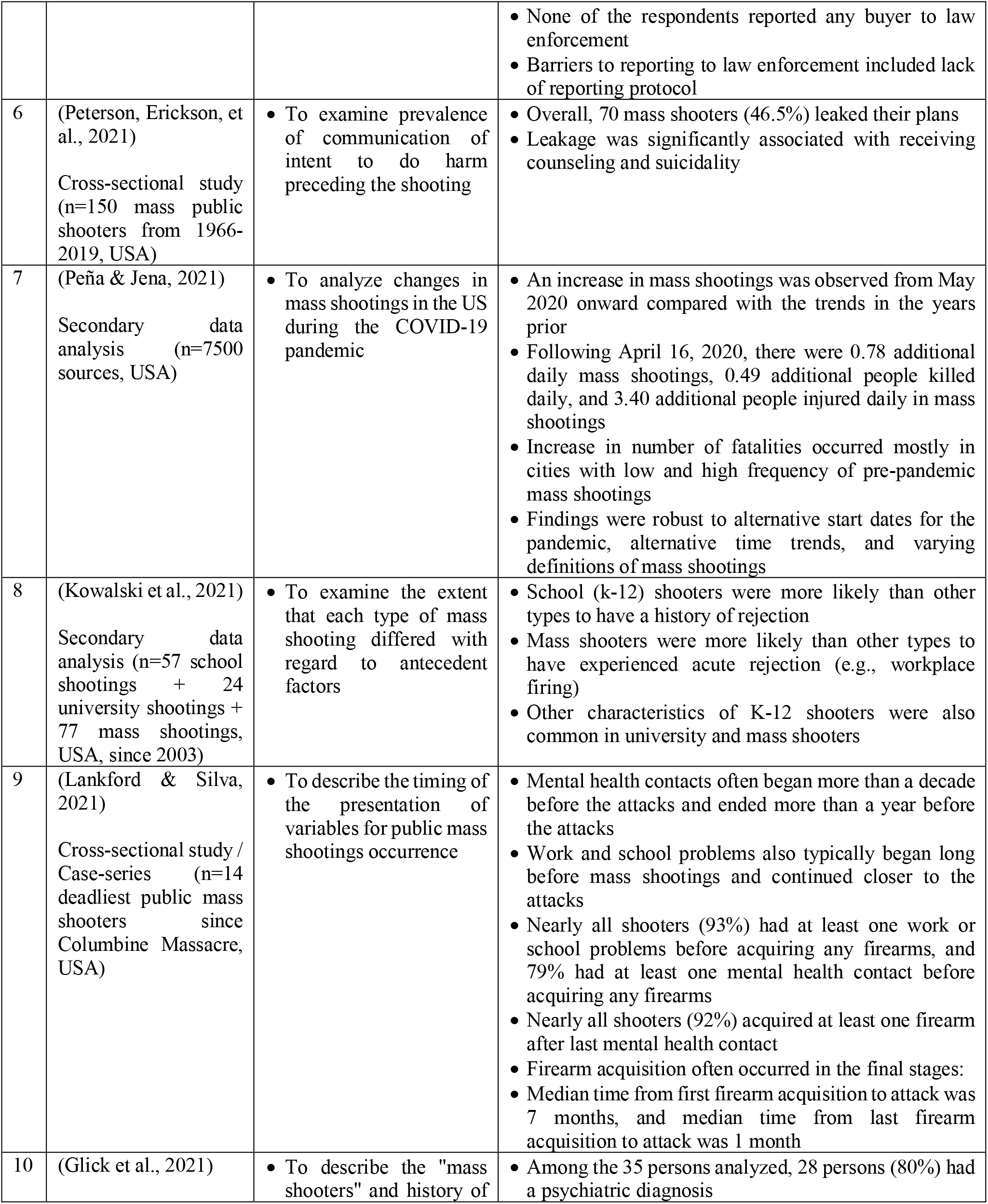
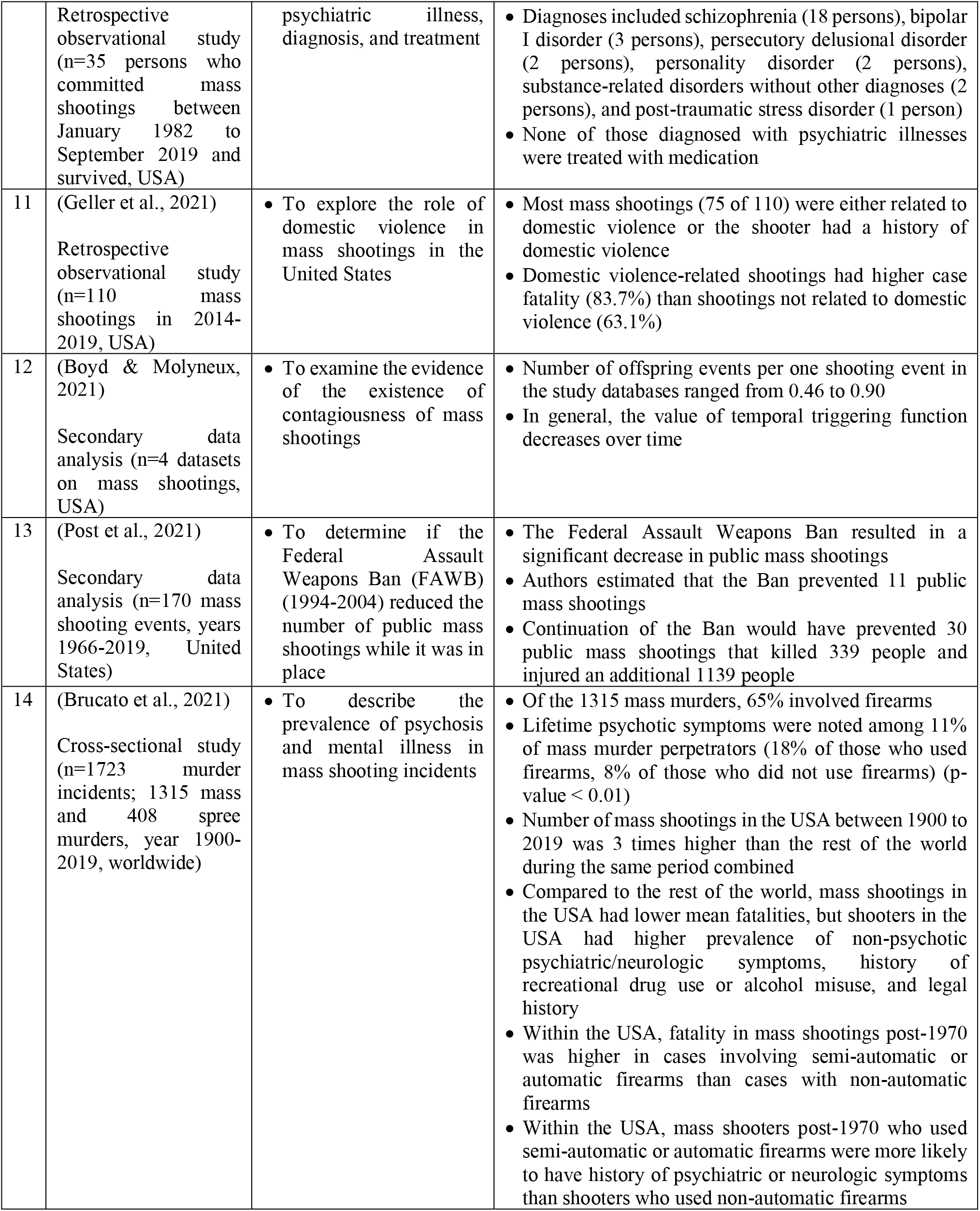
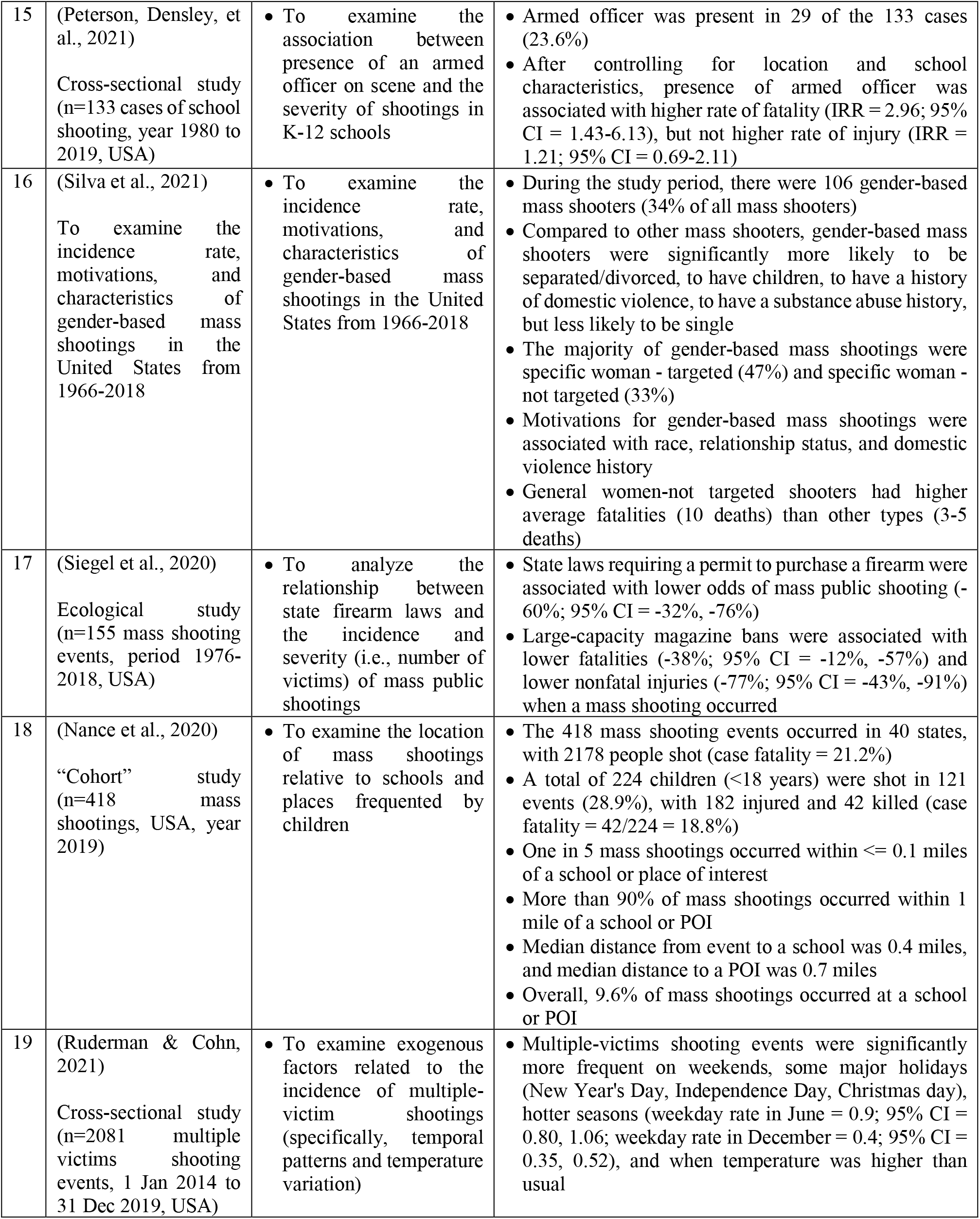
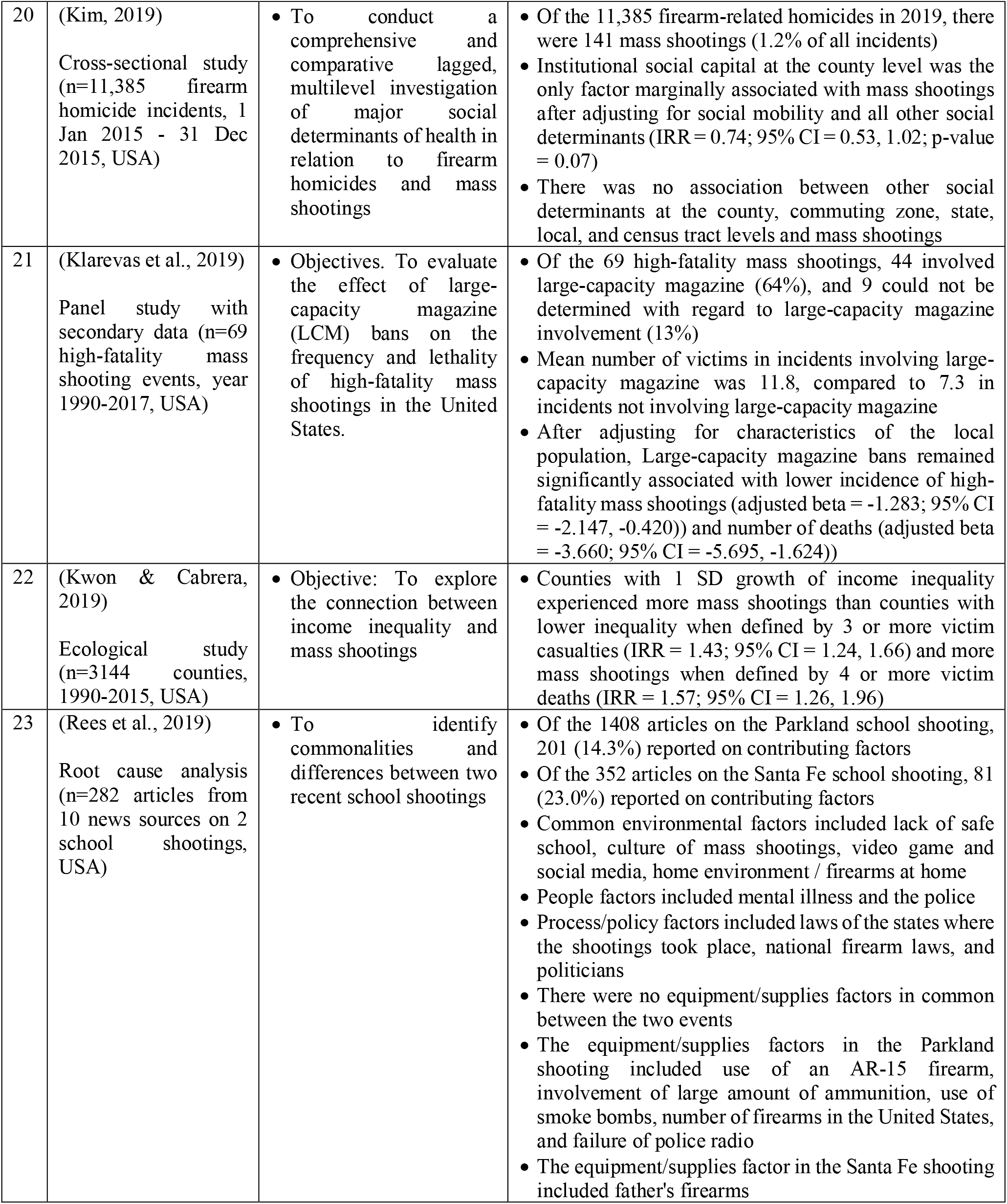
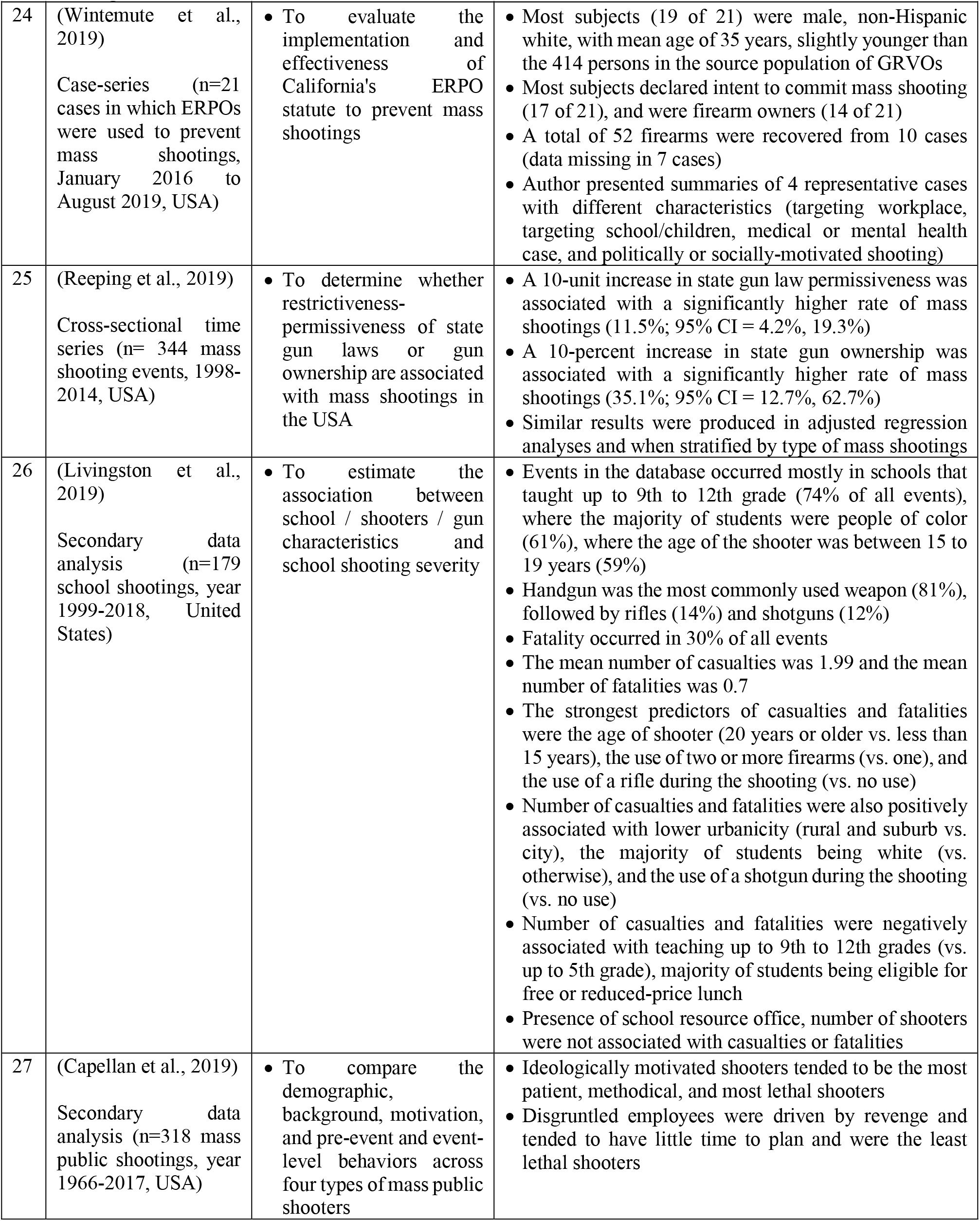
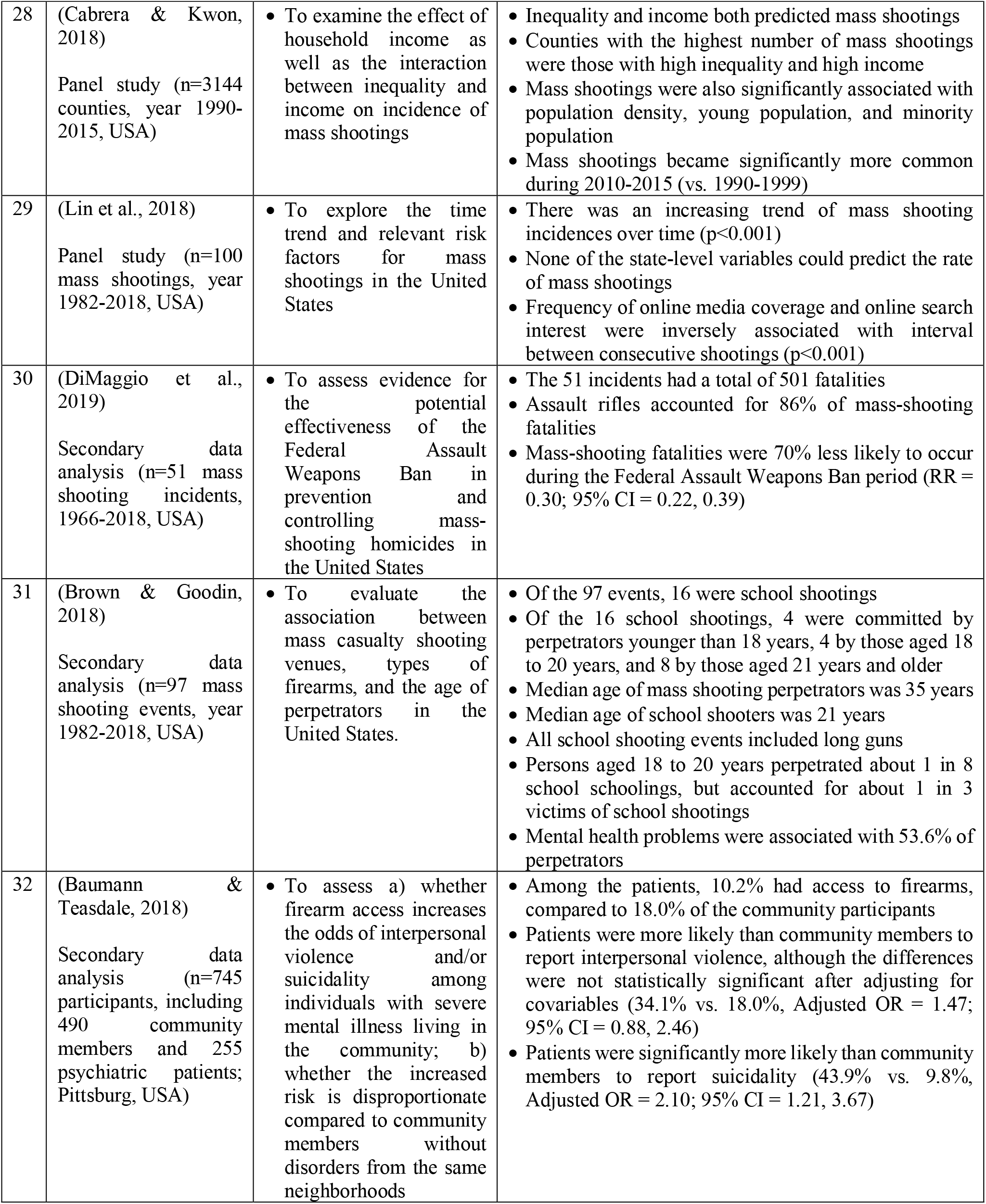
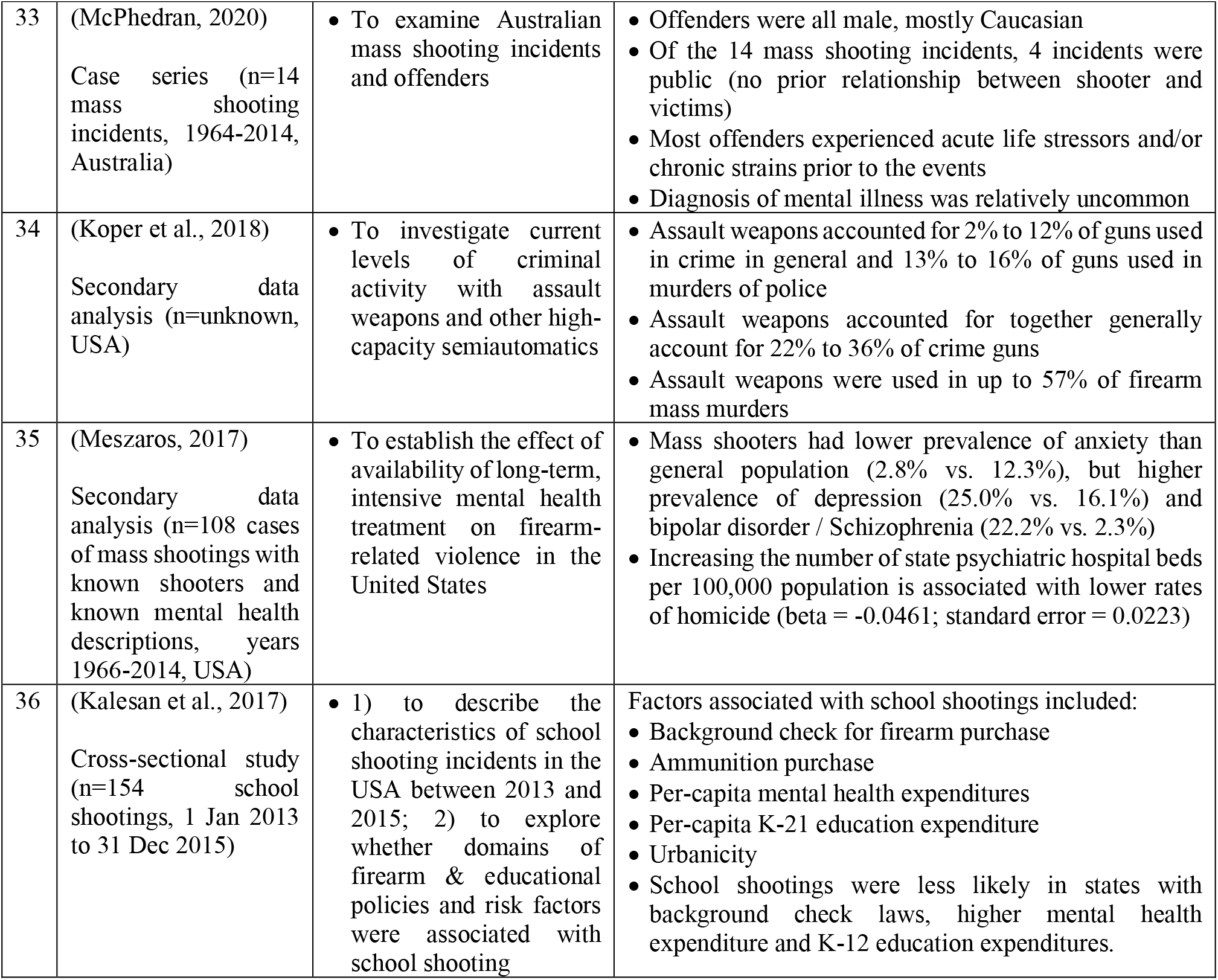
Summary of studies pertaining to perpetration of mass shooting and their determinants (n=36 studies)

| ID | Author and year; Study Design, Population, and Setting | Objective(s) | Results |
| --- | --- | --- | --- |
| 1 | (Reeping et al., 2022)<br>Secondary data analysis (n=unknown; U.S. states) | To identify factors associated with intentional school shootings | State-level firearm laws permissiveness and rate of gun ownership were associated with higher rates of school shootings and active school shootings |
| 2 | (Silver & Silva, 2022)<br>Secondary data analysis (n=unknown; mass shooters) | To assess the order in which public mass shooters encountered behavioral and experiential factors | Shooters lived in coherent phases<br>There was centrality of family problems and interest in past mass killings in the overall sequence |
| 3 | (Duchesne et al., 2022)<br>Secondary data analysis (n=2736 mass shooting events; United States) | To assess the association between strength of state gun laws and incidence of mass shooting event | The incidence of mass shooting events increased from 272 in 2014 to 626<br>Mean mass shooting incidence in states with weakest gun laws increased from 4.0 per million to 9.7 per million<br>There were no significant differences in number of mass shooting events per year by gun law grade, before and after adjusting for population |
| 4 | (Cerfolio et al., 2022, p. 20)<br>Secondary data analysis (n=55 mass shooters, United States) | To understand the underlying psychiatric, psychosocial, and psychodynamic of mass shootings in the United States | The majority of shooters (87.5%) were not diagnosed or received a misdiagnosis or incorrect treatment for mental illness<br>Most assailants experienced severe estrangement from families, friends, classmates, and themselves<br>Marginalization and estrangement rendered the shooters more vulnerable to the psychiatric illness and led to online radicalization, which fostered violence |
| 5 | (Freitas & Annas, 2022)<br>Cross-sectional study (n=9 gun sellers, Onondaga County, New York state, United States) | To describe the opinions, beliefs, attitudes, and current practices of gun sellers in Onondaga County, NY, in regard to safe gun sales | Investigators sent 72 surveys to gun sellers, 2 were undeliverable, and 9 sellers returned questionnaires (response rate = 9/70 = 12.9%)<br>Sixty-two percent (62.5%) of respondents had prior experience in uniform services, including specialized training in mental or emotional crises and deception detection<br>Sales denial occurred in 25 of 1471 weapons and ammunition sale made by all qualifying respondents (1.7%)<br>Of the 25 details, 15 were National Instant Criminal Background Check System (NICS) denials, and 10 were personal denials.<br>Reasons for personal denials included "interest in illegal fittings", "information revealed during conversations", and "lack of basic knowledge of using the firearm looking to purchase" |

| ID | Author and year; Study Population, Design, and Setting | Objective(s) | Results |
| --- | --- | --- | --- |
|  |  |  | • None of the respondents reported any buyer to law enforcement<br>• Barriers to reporting to law enforcement included lack of reporting protocol |
| 6 | (Peterson, Erickson, et al., 2021)<br>Cross-sectional study (n=150 mass public shooters from 1966-2019, USA) | • To examine prevalence of communication of intent to do harm preceding the shooting | • Overall, 70 mass shooters (46.5%) leaked their plans<br>• Leakage was significantly associated with receiving counseling and suicidality |
| 7 | (Peña & Jena, 2021)<br>Secondary data analysis (n=7500 sources, USA) | • To analyze changes in mass shootings in the US during the COVID-19 pandemic | • An increase in mass shootings was observed from May 2020 onward compared with the trends in the years prior<br>• Following April 16, 2020, there were 0.78 additional daily mass shootings, 0.49 additional people killed daily, and 3.40 additional people injured daily in mass shootings<br>• Increase in number of fatalities occurred mostly in cities with low and high frequency of pre-pandemic mass shootings<br>• Findings were robust to alternative start dates for the pandemic, alternative time trends, and varying definitions of mass shootings |
| 8 | (Kowalski et al., 2021)<br>Secondary data analysis (n=57 school shootings + 24 university shootings + 77 mass shootings, USA, since 2003) | • To examine the extent that each type of mass shooting differed with regard to antecedent factors | • School (k-12) shooters were more likely than other types to have a history of rejection<br>• Mass shooters were more likely than other types to have experienced acute rejection (e.g., workplace firing)<br>• Other characteristics of K-12 shooters were also common in university and mass shooters |
| 9 | (Lankford & Silva, 2021)<br>Cross-sectional study / Case-series (n=14 deadliest public mass shooters since Columbine Massacre, USA) | • To describe the timing of the presentation of variables for public mass shootings occurrence | • Mental health contacts often began more than a decade before the attacks and ended more than a year before the attacks<br>• Work and school problems also typically began long before mass shootings and continued closer to the attacks<br>• Nearly all shooters (93%) had at least one work or school problems before acquiring any firearms, and 79% had at least one mental health contact before acquiring any firearms<br>• Nearly all shooters (92%) acquired at least one firearm after last mental health contact<br>• Firearm acquisition often occurred in the final stages:<br>• Median time from first firearm acquisition to attack was 7 months, and median time from last firearm acquisition to attack was 1 month |
| 10 | (Glick et al., 2021) | • To describe the "mass shooters" and history of | • Among the 35 persons analyzed, 28 persons (80%) had a psychiatric diagnosis |
|  | Retrospective observational study (n=35 persons who committed mass shootings between January 1982 to September 2019 and survived, USA) | psychiatric illness, diagnosis, and treatment | Diagnoses included schizophrenia (18 persons), bipolar I disorder (3 persons), persecutory delusional disorder (2 persons), personality disorder (2 persons), substance-related disorders without other diagnoses (2 persons), and post-traumatic stress disorder (1 person)<br>None of those diagnosed with psychiatric illnesses were treated with medication |
| 11 | (Geller et al., 2021)<br>Retrospective observational study (n=110 mass shootings in 2014-2019, USA) | To explore the role of domestic violence in mass shootings in the United States | Most mass shootings (75 of 110) were either related to domestic violence or the shooter had a history of domestic violence<br>Domestic violence-related shootings had higher case fatality (83.7%) than shootings not related to domestic violence (63.1%) |
| 12 | (Boyd & Molyneux, 2021)<br>Secondary data analysis (n=4 datasets on mass shootings, USA) | To examine the evidence of the existence of contagiousness of mass shootings | Number of offspring events per one shooting event in the study databases ranged from 0.46 to 0.90<br>In general, the value of temporal triggering function decreases over time |
| 13 | (Post et al., 2021)<br>Secondary data analysis (n=170 mass shooting events, years 1966-2019, United States) | To determine if the Federal Assault Weapons Ban (FAWB) (1994-2004) reduced the number of public mass shootings while it was in place | The Federal Assault Weapons Ban resulted in a significant decrease in public mass shootings<br>Authors estimated that the Ban prevented 11 public mass shootings<br>Continuation of the Ban would have prevented 30 public mass shootings that killed 339 people and injured an additional 1139 people |
| 14 | (Brucato et al., 2021)<br>Cross-sectional study (n=1723 murder incidents; 1315 mass and 408 spree murders, year 1900-2019, worldwide) | To describe the prevalence of psychosis and mental illness in mass shooting incidents | Of the 1315 mass murders, 65% involved firearms<br>Lifetime psychotic symptoms were noted among 11% of mass murder perpetrators (18% of those who used firearms, 8% of those who did not use firearms) (p-value < 0.01)<br>Number of mass shootings in the USA between 1900 to 2019 was 3 times higher than the rest of the world during the same period combined<br>Compared to the rest of the world, mass shootings in the USA had lower mean fatalities, but shooters in the USA had higher prevalence of non-psychotic psychiatric/neurologic symptoms, history of recreational drug use or alcohol misuse, and legal history<br>Within the USA, fatality in mass shootings post-1970 was higher in cases involving semi-automatic or automatic firearms than cases with non-automatic firearms<br>Within the USA, mass shooters post-1970 who used semi-automatic or automatic firearms were more likely to have history of psychiatric or neurologic symptoms than shooters who used non-automatic firearms |
| 15 | (Peterson, Densley, et al., 2021)<br>Cross-sectional study (n=133 cases of school shooting, year 1980 to 2019, USA) | To examine the association between presence of an armed officer on scene and the severity of shootings in K-12 schools | Armed officer was present in 29 of the 133 cases (23.6%)<br>After controlling for location and school characteristics, presence of armed officer was associated with higher rate of fatality (IRR = 2.96; 95% CI = 1.43-6.13), but not higher rate of injury (IRR = 1.21; 95% CI = 0.69-2.11) |
| 16 | (Silva et al., 2021)<br>To examine the incidence rate, motivations, and characteristics of gender-based mass shootings in the United States from 1966-2018 | To examine the incidence rate, motivations, and characteristics of gender-based mass shootings in the United States from 1966-2018 | During the study period, there were 106 gender-based mass shooters (34% of all mass shooters)<br>Compared to other mass shooters, gender-based mass shooters were significantly more likely to be separated/divorced, to have children, to have a history of domestic violence, to have a substance abuse history, but less likely to be single<br>The majority of gender-based mass shootings were specific woman - targeted (47%) and specific woman - not targeted (33%)<br>Motivations for gender-based mass shootings were associated with race, relationship status, and domestic violence history<br>General women-not targeted shooters had higher average fatalities (10 deaths) than other types (3-5 deaths) |
| 17 | (Siegel et al., 2020)<br>Ecological study (n=155 mass shooting events, period 1976-2018, USA) | To analyze the relationship between state firearm laws and the incidence and severity (i.e., number of victims) of mass public shootings | State laws requiring a permit to purchase a firearm were associated with lower odds of mass public shooting (-60%; 95% CI = -32%, -76%)<br>Large-capacity magazine bans were associated with lower fatalities (-38%; 95% CI = -12%, -57%) and lower nonfatal injuries (-77%; 95% CI = -43%, -91%) when a mass shooting occurred |
| 18 | (Nance et al., 2020)<br>“Cohort” study (n=418 mass shootings, USA, year 2019) | To examine the location of mass shootings relative to schools and places frequented by children | The 418 mass shooting events occurred in 40 states, with 2178 people shot (case fatality = 21.2%)<br>A total of 224 children (<18 years) were shot in 121 events (28.9%), with 182 injured and 42 killed (case fatality = 42/224 = 18.8%)<br>One in 5 mass shootings occurred within <= 0.1 miles of a school or place of interest<br>More than 90% of mass shootings occurred within 1 mile of a school or POI<br>Median distance from event to a school was 0.4 miles, and median distance to a POI was 0.7 miles<br>Overall, 9.6% of mass shootings occurred at a school or POI |
| 19 | (Ruderman & Cohn, 2021)<br>Cross-sectional study (n=2081 multiple victims shooting events, 1 Jan 2014 to 31 Dec 2019, USA) | To examine exogenous factors related to the incidence of multiple-victim shootings (specifically, temporal patterns and temperature variation) | Multiple-victims shooting events were significantly more frequent on weekends, some major holidays (New Year's Day, Independence Day, Christmas day), hotter seasons (weekday rate in June = 0.9; 95% CI = 0.80, 1.06; weekday rate in December = 0.4; 95% CI = 0.35, 0.52), and when temperature was higher than usual |
| 20 | (Kim, 2019)<br>Cross-sectional study (n=11,385 firearm homicide incidents, 1 Jan 2015 - 31 Dec 2015, USA) | To conduct a comprehensive and comparative lagged, multilevel investigation of major social determinants of health in relation to firearm homicides and mass shootings | Of the 11,385 firearm-related homicides in 2019, there were 141 mass shootings (1.2% of all incidents)<br>Institutional social capital at the county level was the only factor marginally associated with mass shootings after adjusting for social mobility and all other social determinants (IRR = 0.74; 95% CI = 0.53, 1.02; p-value = 0.07)<br>There was no association between other social determinants at the county, commuting zone, state, local, and census tract levels and mass shootings |
| 21 | (Klarevas et al., 2019)<br>Panel study with secondary data (n=69 high-fatality mass shooting events, year 1990-2017, USA) | Objectives. To evaluate the effect of large-capacity magazine (LCM) bans on the frequency and lethality of high-fatality mass shootings in the United States. | Of the 69 high-fatality mass shootings, 44 involved large-capacity magazine (64%), and 9 could not be determined with regard to large-capacity magazine involvement (13%)<br>Mean number of victims in incidents involving large-capacity magazine was 11.8, compared to 7.3 in incidents not involving large-capacity magazine<br>After adjusting for characteristics of the local population, Large-capacity magazine bans remained significantly associated with lower incidence of high-fatality mass shootings (adjusted beta = -1.283; 95% CI = -2.147, -0.420)) and number of deaths (adjusted beta = -3.660; 95% CI = -5.695, -1.624)) |
| 22 | (Kwon & Cabrera, 2019)<br>Ecological study (n=3144 counties, 1990-2015, USA) | Objective: To explore the connection between income inequality and mass shootings | Counties with 1 SD growth of income inequality experienced more mass shootings than counties with lower inequality when defined by 3 or more victim casualties (IRR = 1.43; 95% CI = 1.24, 1.66) and more mass shootings when defined by 4 or more victim deaths (IRR = 1.57; 95% CI = 1.26, 1.96) |
| 23 | (Rees et al., 2019)<br>Root cause analysis (n=282 articles from 10 news sources on 2 school shootings, USA) | To identify commonalities and differences between two recent school shootings | Of the 1408 articles on the Parkland school shooting, 201 (14.3%) reported on contributing factors<br>Of the 352 articles on the Santa Fe school shooting, 81 (23.0%) reported on contributing factors<br>Common environmental factors included lack of safe school, culture of mass shootings, video game and social media, home environment / firearms at home<br>People factors included mental illness and the police<br>Process/policy factors included laws of the states where the shootings took place, national firearm laws, and politicians<br>There were no equipment/supplies factors in common between the two events<br>The equipment/supplies factors in the Parkland shooting included use of an AR-15 firearm, involvement of large amount of ammunition, use of smoke bombs, number of firearms in the United States, and failure of police radio<br>The equipment/supplies factor in the Santa Fe shooting included father's firearms |
| 24 | (Wintemute et al., 2019)<br>Case-series (n=21 cases in which ERPOs were used to prevent mass shootings, January 2016 to August 2019, USA) | To evaluate the implementation and effectiveness of California's ERPO statute to prevent mass shootings | Most subjects (19 of 21) were male, non-Hispanic white, with mean age of 35 years, slightly younger than the 414 persons in the source population of GRVOs<br>Most subjects declared intent to commit mass shooting (17 of 21), and were firearm owners (14 of 21)<br>A total of 52 firearms were recovered from 10 cases (data missing in 7 cases)<br>Author presented summaries of 4 representative cases with different characteristics (targeting workplace, targeting school/children, medical or mental health case, and politically or socially-motivated shooting) |
| 25 | (Reeping et al., 2019)<br>Cross-sectional time series (n= 344 mass shooting events, 1998-2014, USA) | To determine whether restrictiveness-permissiveness of state gun laws or gun ownership are associated with mass shootings in the USA | A 10-unit increase in state gun law permissiveness was associated with a significantly higher rate of mass shootings (11.5%; 95% CI = 4.2%, 19.3%)<br>A 10-percent increase in state gun ownership was associated with a significantly higher rate of mass shootings (35.1%; 95% CI = 12.7%, 62.7%)<br>Similar results were produced in adjusted regression analyses and when stratified by type of mass shootings |
| 26 | (Livingston et al., 2019)<br>Secondary data analysis (n=179 school shootings, year 1999-2018, United States) | To estimate the association between school / shooters / gun characteristics and school shooting severity | Events in the database occurred mostly in schools that taught up to 9th to 12th grade (74% of all events), where the majority of students were people of color (61%), where the age of the shooter was between 15 to 19 years (59%)<br>Handgun was the most commonly used weapon (81%), followed by rifles (14%) and shotguns (12%)<br>Fatality occurred in 30% of all events<br>The mean number of casualties was 1.99 and the mean number of fatalities was 0.7<br>The strongest predictors of casualties and fatalities were the age of shooter (20 years or older vs. less than 15 years), the use of two or more firearms (vs. one), and the use of a rifle during the shooting (vs. no use)<br>Number of casualties and fatalities were also positively associated with lower urbanicity (rural and suburb vs. city), the majority of students being white (vs. otherwise), and the use of a shotgun during the shooting (vs. no use)<br>Number of casualties and fatalities were negatively associated with teaching up to 9th to 12th grades (vs. up to 5th grade), majority of students being eligible for free or reduced-price lunch<br>Presence of school resource office, number of shooters were not associated with casualties or fatalities |
| 27 | (Capellan et al., 2019)<br>Secondary data analysis (n=318 mass public shootings, year 1966-2017, USA) | To compare the demographic, background, motivation, and pre-event and event-level behaviors across four types of mass public shooters | Ideologically motivated shooters tended to be the most patient, methodical, and most lethal shooters<br>Disgruntled employees were driven by revenge and tended to have little time to plan and were the least lethal shooters |
| 28 | (Cabrera & Kwon, 2018)<br>Panel study (n=3144 counties, year 1990-2015, USA) | To examine the effect of household income as well as the interaction between inequality and income on incidence of mass shootings | Inequality and income both predicted mass shootings<br>Counties with the highest number of mass shootings were those with high inequality and high income<br>Mass shootings were also significantly associated with population density, young population, and minority population<br>Mass shootings became significantly more common during 2010-2015 (vs. 1990-1999) |
| 29 | (Lin et al., 2018)<br>Panel study (n=100 mass shootings, year 1982-2018, USA) | To explore the time trend and relevant risk factors for mass shootings in the United States | There was an increasing trend of mass shooting incidences over time ($p < 0.001$)<br>None of the state-level variables could predict the rate of mass shootings<br>Frequency of online media coverage and online search interest were inversely associated with interval between consecutive shootings ($p < 0.001$) |
| 30 | (DiMaggio et al., 2019)<br>Secondary data analysis (n=51 mass shooting incidents, 1966-2018, USA) | To assess evidence for the potential effectiveness of the Federal Assault Weapons Ban in prevention and controlling mass-shooting homicides in the United States | The 51 incidents had a total of 501 fatalities<br>Assault rifles accounted for 86% of mass-shooting fatalities<br>Mass-shooting fatalities were 70% less likely to occur during the Federal Assault Weapons Ban period (RR = 0.30; 95% CI = 0.22, 0.39) |
| 31 | (Brown & Goodin, 2018)<br>Secondary data analysis (n=97 mass shooting events, year 1982-2018, USA) | To evaluate the association between mass casualty shooting venues, types of firearms, and the age of perpetrators in the United States. | Of the 97 events, 16 were school shootings<br>Of the 16 school shootings, 4 were committed by perpetrators younger than 18 years, 4 by those aged 18 to 20 years, and 8 by those aged 21 years and older<br>Median age of mass shooting perpetrators was 35 years<br>Median age of school shooters was 21 years<br>All school shooting events included long guns<br>Persons aged 18 to 20 years perpetrated about 1 in 8 school shootings, but accounted for about 1 in 3 victims of school shootings<br>Mental health problems were associated with 53.6% of perpetrators |
| 32 | (Baumann & Teasdale, 2018)<br>Secondary data analysis (n=745 participants, including 490 community members and 255 psychiatric patients; Pittsburg, USA) | To assess a) whether firearm access increases the odds of interpersonal violence and/or suicidality among individuals with severe mental illness living in the community; b) whether the increased risk is disproportionate compared to community members without disorders from the same neighborhoods | Among the patients, 10.2% had access to firearms, compared to 18.0% of the community participants<br>Patients were more likely than community members to report interpersonal violence, although the differences were not statistically significant after adjusting for covariables (34.1% vs. 18.0%, Adjusted OR = 1.47; 95% CI = 0.88, 2.46)<br>Patients were significantly more likely than community members to report suicidality (43.9% vs. 9.8%, Adjusted OR = 2.10; 95% CI = 1.21, 3.67) |
| 33 | (McPhedran, 2020)<br>Case series (n=14 mass shooting incidents, 1964-2014, Australia) | To examine Australian mass shooting incidents and offenders | Offenders were all male, mostly Caucasian<br>Of the 14 mass shooting incidents, 4 incidents were public (no prior relationship between shooter and victims)<br>Most offenders experienced acute life stressors and/or chronic strains prior to the events<br>Diagnosis of mental illness was relatively uncommon |
| 34 | (Koper et al., 2018)<br>Secondary data analysis (n=unknown, USA) | To investigate current levels of criminal activity with assault weapons and other high-capacity semiautomatics | Assault weapons accounted for 2% to 12% of guns used in crime in general and 13% to 16% of guns used in murders of police<br>Assault weapons accounted for together generally account for 22% to 36% of crime guns<br>Assault weapons were used in up to 57% of firearm mass murders |
| 35 | (Meszaros, 2017)<br>Secondary data analysis (n=108 cases of mass shootings with known shooters and known mental health descriptions, years 1966-2014, USA) | To establish the effect of availability of long-term, intensive mental health treatment on firearm-related violence in the United States | Mass shooters had lower prevalence of anxiety than general population (2.8% vs. 12.3%), but higher prevalence of depression (25.0% vs. 16.1%) and bipolar disorder / Schizophrenia (22.2% vs. 2.3%)<br>Increasing the number of state psychiatric hospital beds per 100,000 population is associated with lower rates of homicide (beta = -0.0461; standard error = 0.0223) |
| 36 | (Kalesan et al., 2017)<br>Cross-sectional study (n=154 school shootings, 1 Jan 2013 to 31 Dec 2015) | 1) to describe the characteristics of school shooting incidents in the USA between 2013 and 2015; 2) to explore whether domains of firearm & educational policies and risk factors were associated with school shooting | Factors associated with school shootings included:<br>Background check for firearm purchase<br>Ammunition purchase<br>Per-capita mental health expenditures<br>Per-capita K-21 education expenditure<br>Urbanicity<br>School shootings were less likely in states with background check laws, higher mental health expenditure and K-12 education expenditures. |

**Table 2.** Factors associated with perpetration of mass shootings in the social ecological model

| Level description of factors | Study reference number |  |  |
| --- | --- | --- | --- |
|  | Positive | Negative | Neutral |
| ***Intrapersonal factors*** |  |  |  |
| *Health-related issues* |  |  |  |
| Mental illness (any type) | [4], [9], [10], [14], [15], [23], [31], [35] | [35] | [32] |
| *Psychology / Behaviors* |  |  |  |
| Interest in past mass killings | [2] |  |  |
| Communication of Intent | [6], [24] |  |  |
| Estrangement from self | [4] |  |  |
| Online radicalization | [4] |  |  |
| Substance abuse | [16] |  |  |
| Video game and social media use | [23] |  |  |
| Gun ownership | [1], [9] |  |  |
| *Personal Experience* |  |  |  |
| Experience of acute life stressor | [33] |  |  |
| ***Interpersonal factors*** |  |  |  |
| Family problems | [2] |  |  |
| Estrangement (from families, friends, classmates) | [4] |  |  |
| History of domestic violence | [11], [16] |  |  |
| History of rejection | [8] |  |  |
| Being single |  | [16] |  |
| Having children | [16] |  |  |
| Being separated or divorced | [16] |  |  |
| Home environment contains firearms | [23] |  |  |
| ***Organizational factors*** |  |  |  |
| Work and school problems | [9] |  |  |
| ***Community factors*** |  |  |  |
| *Gun ownership and gun control* |  |  |  |
| State gun ownership | [25] |  |  |
| Community's access to assault weapons | [34] |  |  |
| Ammunition purchase | [36] |  |  |
| NICS sales denial |  | [5] |  |
| *Social-cultural factors* |  |  |  |
| Culture of mass shootings | [23] |  |  |
| Online media coverage | [29] |  |  |
| Institutional social capital |  | [20] |  |
| *Contextual factors* |  |  |  |
| Time (more recent years) | [28],[29] |  |  |
|  | Positive | Negative | Neutral |
| Time (COVID-19 pandemic onset) | [7] |  |  |
| Time (major public holidays or hotter seasons) | [19] |  |  |
| Place (Urbanicity) | [26] |  |  |
| Place (near school or places of interest to children) | [18] |  |  |
| Place (occurrence of previous shooting in area) |  | [12] |  |
| Demographics (Income inequality) | [22],[28] |  |  |
| Demographics (Population density) | [28] |  |  |
| Demographics (Population median age) |  | [28] |  |
| Others (Lack of safe school) | [23] |  |  |
| Others (Number of state psychiatric hospitals) |  | [35] |  |
| ***Policy factors*** |  |  |  |
| *Gun ownership and gun control* |  |  |  |
| State firearm laws (permissive) | [1], [25] |  | [3] |
| Background check for firearm purchase |  | [36] |  |
| Requirement of permit to purchase firearms |  | [17] |  |
| Federal Assault Weapons Ban (FAWB) |  | [13] |  |
| Large-capacity magazine ban |  | [21] |  |
| *Legal / Social / Health interventions* |  |  |  |
| Emergency Protection Order (ERPO / GRVO) |  | [24] |  |
| Per-capita mental health and K-12 education expenditures |  | [36] |  |

**Table 3.** Factors associated with fatalities and injuries at mass shooting events in the social ecological model

| Level description of factors | Study reference number |  |  |
| --- | --- | --- | --- |
|  | Positive | Negative | Neutral |
| ***Intrapersonal factors*** |  |  |  |
| *Firearms-related factors* |  |  |  |
| Use of semi-automatic or automatic firearms | [14] |  |  |
| Use of a rifle during shooting | [26],[31] |  |  |
| Use of a shotgun during shooting | [26] |  |  |
| Use of two or more firearms | [26] |  |  |
| *Psychology / Behaviors* |  |  |  |
| Targeting women in general (no specific person as target) | [16] |  |  |
| Being ideologically motivated and methodical | [27] |  |  |
| Being driven by revenge as an employee |  | [27] |  |
| *Demographics* |  |  |  |
| Older age of school shooter | [26],[31] |  |  |
| ***Interpersonal factors*** |  |  |  |
| History of domestic violence | [11] |  |  |
| ***Organizational factors*** |  |  |  |
| Presence of armed officer at school | [15] |  | [15],[26] |
| School teaching up to 9th to 12th grade (vs. up to younger grades) | [26] |  |  |
| School with students eligible for reduced-price lunch | [26] |  |  |
| ***Community factors*** |  |  |  |
| *Contextual factors* |  |  |  |
| Place (Urbanicity) |  | [26] |  |
| Demographic (Caucasian-majority) | [26] |  |  |
| ***Policy factors*** |  |  |  |
| *Gun ownership and gun control* |  |  |  |
| Federal Assault Weapons Ban (FAWB) |  | [30] |  |
| Large-capacity magazine ban |  | [17],[21] |  |

At the community level, gun ownership, access to assault weapons and ammunition were all positively associated with perpetration of mass shooting. Other community-level factors included the presence of a culture of mass shooting and online media coverage of mass shooting. Contexts including time, place, and demographics were also associated with perpetration of mass shootings. Mass shootings have become more common in the 2010s compared to the 2000s and the 1990s, particularly during the COVID-19 pandemic, during major public holidays, and in hotter seasons. Mass shootings were more common in urban areas, generally occurred within one mile of a school or places of interest frequented by children, and more common in areas with higher income inequality and higher population density. However, mass shootings were less common in places with higher population median age, and in states with higher number of psychiatric hospitals. At the policy level, permissive state firearm laws were either neutral or positively associated with perpetration of mass shooting. On the other hand, gun control measures (background check, permit requirement, Federal Assault Weapons Ban, and large-capacity magazine ban) were all associated with lower perpetration of mass shootings. Other measures such as emergency protection order (ERPO) and mental health expenditures per capita also showed a preventive relationship with perpetration of mass shootings.

### Factors associated with mass shooting injuries and fatalities

At the intrapersonal and interpersonal levels, firearm characteristics were associated with injuries and fatalities. Although handguns were the most common type of firearm used in mass shootings, shootings that involved semi-automatic or automatic firearms had higher mean fatality than shootings that involved only handguns. Number of casualties at mass shootings was also positively associated with use of two or more firearms. Gender-based mass shooters who targeted women, and ideologically-motivated shooters, tended to be more patient and lethal. Disgruntled employees who perpetrated mass shootings generally had little or no time to plan and tended to be less lethal. School shooters who were older (aged 18-20 years) harmed more persons in each attack than school-aged shooters. Mass shootings related to domestic violence were more lethal than mass shootings not related to domestic violence. At the organizational and community levels, most data pertained to school shootings. Mass shootings at high school tended to have more casualties than those perpetrated at elementary or middle schools. Shooting injuries and fatalities were also more common at schools with students eligible for reduced-price lunch. On the other hand, presence of armed officer at school either had no association with casualties at mass shooting events, or was associated with higher (not lower) casualties. Mass shootings were less common in urban schools and more common in schools with a Caucasian-majority student body. At the policy level, gun control measures including Federal Assault Weapons Ban and large-capacity magazine ban were associated with lower fatality and nonfatal injuries at mass shootings.

## DISCUSSION

In this study, the author systematically reviewed factors associated with perpetration of mass shooting events, and factors associated with injuries and fatalities at mass shooting events. The author then identified intrapersonal, interpersonal, organization, community, and policy factors associated with perpetration and casualties of mass shootings events. The findings of this study should be of interest to stakeholders, particularly during this period where the frequency of event is ever-increasing in the United States. As the author used the social ecological model to describe the factors associated with the outcomes, the discussion of the study findings should also be disaggregated according to the levels of the model.

### Intrapersonal level

History of mental illness and lack of or discontinuation of treatment were commonly-mentioned factors. Other reports mentioned signs of devolution of the shooter’s behaviors to gravitate towards mass shootings, until finally firearms were acquired and the mass shooting event occurred. The event would be more fatal had it involved any type of semi-automatic firearm or shotgun. However, one must refrain from using these findings to stigmatize those with mental illnesses or estrangement behaviors, as practically all of nearly 53 million US adults with mental illness (National Institute of Mental Health, 2022) do not have plan to harm others, especially not on a massive scale. The study findings should only be used as an attempt to understand the phenomenon in a complete manner.

### Interpersonal level

Problems in interpersonal relationship seemed to be a recurring theme for perpetration of mass shootings. In particular, family problems (including domestic violence, separation or divorce) and rejection from others were common among mass shooters. Domestic violence was also associated with lethality of mass shootings. Considering that mental health problems are associated with family/relationship issues (Silver & Silva, 2022), and that family/relationship issues can lead to mental health problems (Gibb et al., 2011), it is not possible to ascertain whether a variable was a risk factor or the pathway variable between another risk factor and the outcomes.

### Organization level

Perpetration of mass shooting was associated with work or school problem. Further investigation is recommended on the extent that management practice or policy of the workplace or the school contributed to work or school problem, which in turn contributed to the perpetration of mass shooting. Concerningly, presence of armed police officers at schools had an opposite effect to what was intended: no reduction or actual increase in casualties at school shooting events. Data from another source (Justice Policy Institute, 2021) showed that weapons-related arrests accounted for only 0.1% to 0.2% of all arrests made by school resource officers (school police officers), and the majority were for disorderly conduct that funneled students unnecessarily into the juvenile justice system. Given the consistency of the evidence, relevant stakeholders should reconsider the presence of armed guards or police officers in schools.

### Community level

Factors at the community level appeared coherent and complementary to the factors identified at other levels. There were variations in mass shooting with regards to time, place, and demographics of the shooting’s location. Associations between perpetration and casualties of mass shootings were also clear: states with higher gun ownership and access to assault weapons and ammunition had higher incidents of mass shootings than states that were less permissive of firearms. With such clear patterns of association, local government and community leaders should consider increasing gun control where possible.

### Policy level

There were clear patterns of association between gun control measures and perpetration, injuries, and fatalities of mass shootings. In particular, the Federal Assault Weapons Ban (FAWB) and large-capacity magazine ban had negative associations with both perpetration and casualties of mass shootings. Considering that more lethal shootings tended to involve the use of assault weapons with higher number of ammunitions (Brown & Goodin, 2018; Livingston et al., 2019), these policies seemed to directly address these issues. Similarly, other preventive measures such as the emergency protection order and mental health expenditures were also negatively associated with perpetration of mass shootings, suggesting that other measures should also be adopted concurrently with gun control.

A number of limitations should be considered in the findings of this study. Firstly, this review was written in relative haste in response to high-profile incidents of mass shootings in the United States and included studies published on PubMed since 2017, an arbitrary time frame. More comprehensive systematic reviews could have yielded different results. Secondly, the level in the social ecological model to which each potential determinant belonged was relatively subjective and based on the author’s own judgement. Findings from other authors could have been different from those presented by the author.

## CONCLUSION

In this study, the author reviewed factors associated with perpetration and casualty of mass shootings using the social ecological model. Although the author identified factors at various levels, easy access to high-power firearms and lack of gun control were associated with both mass shooting perpetration and casualty. Caveats including a limited time frame, limited sources of publications, and subjectivity in building social ecological model should be considered in the interpretation of the study findings.

## Data Availability

All data produced in the present study are available upon reasonable request to the authors

## Notes

### Competing Interest Statement

The authors have declared no competing interest.

### Funding Statement

This study did not receive any funding

## REFERENCES

Baumann, M. L., & Teasdale, B. (2018). Severe mental illness and firearm access: Is violence really the danger? International Journal of Law and Psychiatry, 56, 44–49. https://doi.org/10.1016/j.ijlp.2017.11.003

Boyd, P., & Molyneux, J. (2021). Assessing the contagiousness of mass shootings with nonparametric Hawkes processes. PloS One, 16(3), e0248437.#x2013;e0248437. PubMed. https://doi.org/10.1371/journal.pone.0248437

Brown, J. D., & Goodin, A. J. (2018). Mass Casualty Shooting Venues, Types of Firearms, and Age of Perpetrators in the United States, 1982-2018. American Journal of Public Health, 108(10), 1385– 1387. PubMed. https://doi.org/10.2105/AJPH.2018.304584

Brucato, G., Appelbaum, P. S., Hesson, H., Shea, E. A., Dishy, G., Lee, K., Pia, T., Syed, F., Villalobos, A., Wall, M. M., Lieberman, J. A., & Girgis, R. R. (2021). Psychotic symptoms in mass shootings v. Mass murders not involving firearms: Findings from the Columbia mass murder database. Psychological Medicine, 1–9. https://doi.org/10.1017/S0033291721000076

Cabrera, J. F., & Kwon, R. (2018). Income Inequality, Household Income, and Mass Shooting in the United States. Frontiers in Public Health, 6, 294–294. PubMed. https://doi.org/10.3389/fpubh.2018.00294

Capellan, J. A., Johnson, J., Porter, J. R., & Martin, C. (2019). Disaggregating Mass Public Shootings: A Comparative Analysis of Disgruntled Employee, School, Ideologically motivated, and Rampage Shooters. Journal of Forensic Sciences, 64(3), 814–823. https://doi.org/10.1111/1556-4029.13985

Cerfolio, N. E., Glick, I., Kamis, D., & Laurence, M. (2022). A Retrospective Observational Study of Psychosocial Determinants and Psychiatric Diagnoses of Mass Shooters in the United States. Psychodynamic Psychiatry, 1–16. https://doi.org/10.1521/pdps.2022.50.5.001

DiMaggio, C., Avraham, J., Berry, C., Bukur, M., Feldman, J., Klein, M., Shah, N., Tandon, M., & Frangos, S. (2019). Changes in US mass shooting deaths associated with the 1994-2004 federal assault weapons ban: Analysis of open-source data. The Journal of Trauma and Acute Care Surgery, 86(1), 11–19. https://doi.org/10.1097/TA.0000000000002060

Duchesne, J., Taghavi, S., Toraih, E., Simpson, J. T., & Tatum, D. (2022). State Gun Law Grades and Impact on Mass Shooting Event Incidence: An 8-Year Analysis. Journal of the American College of Surgeons, 234(4), 645–651. https://doi.org/10.1097/XCS.0000000000000118

Freitas, C., & Annas, G. D. (2022). Warning behaviors and leaked intent: Potential new avenues to prevent mass shootings. Journal of Forensic Sciences, 67(1), 275–278. https://doi.org/10.1111/1556-4029.14950

Geller, L. B., Booty, M., & Crifasi, C. K. (2021). The role of domestic violence in fatal mass shootings in the United States, 2014-2019. Injury Epidemiology, 8(1), 38–38. PubMed. https://doi.org/10.1186/s40621-021-00330-0

Gibb, S. J., Fergusson, D. M., & Horwood, L. J. (2011). Relationship separation and mental health problems: Findings from a 30-year longitudinal study. The Australian and New Zealand Journal of Psychiatry, 45(2), 163–169. https://doi.org/10.3109/00048674.2010.529603

Glick, I. D., Cerfolio, N. E., Kamis, D., & Laurence, M. (2021). Domestic Mass Shooters: The Association With Unmedicated and Untreated Psychiatric Illness. Journal of Clinical Psychopharmacology, 41(4). https://journals.lww.com/psychopharmacology/Fulltext/2021/07000/Domestic_Mass_ShootersThe_Association_With.5.aspx

Justice Policy Institute. (2021). Education Under Arrest: The Case Against Police in Schools. https://justicepolicy.org/wp-content/uploads/2022/02/educationunderarrest_fullreport.pdf

Kalesan, B., Lagast, K., Villarreal, M., Pino, E., Fagan, J., & Galea, S. (2017). School shootings during 2013-2015 in the USA. Injury Prevention : Journal of the International Society for Child and Adolescent Injury Prevention, 23(5), 321–327. https://doi.org/10.1136/injuryprev-2016-042162

Karp, A. (2018). Estimating Global Civilian-Held Firearms Numbers (p. 12) [Briefing Paper]. Small Arms Survey. https://web.archive.org/web/20180620231909/http://www.smallarmssurvey.org/fileadmin/docs/T-Briefing-Papers/SAS-BP-Civilian-Firearms-Numbers.pdf

Keneally, M. (2019, August 6). Walmart shooting in El Paso among deadliest gun massacres in US history. https://abcnews.go.com/US/10-deadliest-mass-shootings-modern-us-history/story?id=50234345

Kim, D. (2019). Social determinants of health in relation to firearm-related homicides in the United States: A nationwide multilevel cross-sectional study. PLoS Medicine, 16(12), e1002978.#x2013;e1002978. PubMed. https://doi.org/10.1371/journal.pmed.1002978

Klarevas, L., Conner, A., & Hemenway, D. (2019). The Effect of Large-Capacity Magazine Bans on High-Fatality Mass Shootings, 1990-2017. American Journal of Public Health, 109(12), 1754–1761. PubMed. https://doi.org/10.2105/AJPH.2019.305311

Koper, C. S., Johnson, W. D., Nichols, J. L., Ayers, A., & Mullins, N. (2018). Criminal Use of Assault Weapons and High-Capacity Semiautomatic Firearms: An Updated Examination of Local and National Sources. Journal of Urban Health : Bulletin of the New York Academy of Medicine, 95(3), 313–321. PubMed. https://doi.org/10.1007/s11524-017-0205-7

Kowalski, R. M., Leary, M., Hendley, T., Rubley, K., Chapman, C., Chitty, H., Carroll, H., Cook, A., Richardson, E., Robbins, C., Wells, S., Bourque, L., Oakley, R., Bednar, H., Jones, R., Tolleson, K., Fisher, K., Graham, R., Scarborough, M., … Longacre, M. (2021). K-12, college/university, and mass shootings: Similarities and differences. The Journal of Social Psychology, 161(6), 753–778. https://doi.org/10.1080/00224545.2021.1900047

Kwon, R., & Cabrera, J. F. (2019). Income inequality and mass shootings in the United States. BMC Public Health, 19(1), 1147–1147. PubMed. https://doi.org/10.1186/s12889-019-7490-x

Lankford, A., & Silva, J. R. (2021). The timing of opportunities to prevent mass shootings: A study of mental health contacts, work and school problems, and firearms acquisition. International Review of Psychiatry (Abingdon, England), 33(7), 638–652. https://doi.org/10.1080/09540261.2021.1932440

Lin, P.-I., Fei, L., Barzman, D., & Hossain, M. (2018). What have we learned from the time trend of mass shootings in the U.S.? PloS One, 13(10), e0204722.#x2013;e0204722. PubMed. https://doi.org/10.1371/journal.pone.0204722

Livingston, M. D., Rossheim, M. E., & Hall, K. S. (2019). A Descriptive Analysis of School and School Shooter Characteristics and the Severity of School Shootings in the United States, 1999-2018. The Journal of Adolescent Health : Official Publication of the Society for Adolescent Medicine, 64(6), 797–799. https://doi.org/10.1016/j.jadohealth.2018.12.006

McKinley, J., Traub, A., & Closson, T. (2022, May 14). Gunman Kills 10 at Buffalo Supermarket in Racist Attack. The New York Times. https://www.nytimes.com/live/2022/05/14/nyregion/buffalo-shooting

McLeroy, K. R., Bibeau, D., Steckler, A., & Glanz, K. (1988). An ecological perspective on health promotion programs. Health Education Quarterly, 15(4), 351–377. https://doi.org/10.1177/109019818801500401

McPhedran, S. (2020). Australian Mass Shootings: An Analysis of Incidents and Offenders. Journal of Interpersonal Violence, 35(19–20), 3939–3962. https://doi.org/10.1177/0886260517713226

Meszaros, J. (2017). Falling through the cracks: The decline of mental health care and firearm violence. Journal of Mental Health (Abingdon, England), 26(4), 359–365. https://doi.org/10.1080/09638237.2017.1340608

Nance, M. L., DeSimone, J. D., Lorch, S. A., Passarella, M., Cronin, K. M., Kreinces, J., & Myers, S. R. (2020). Locations of Mass Shootings Relative to Schools and Places Frequented by Children. JAMA Pediatrics, 174(11), 1109–1110. PubMed. https://doi.org/10.1001/jamapediatrics.2020.3371

National Institute of Mental Health. (2022). Mental Illness. https://www.nimh.nih.gov/health/statistics/mental-illness

Peña, P. A., & Jena, A. (2021). Mass Shootings in the US During the COVID-19 Pandemic. JAMA Network Open, 4(9), e2125388.#x2013;e2125388. PubMed. https://doi.org/10.1001/jamanetworkopen.2021.25388

Peterson, J., Densley, J., & Erickson, G. (2021). Presence of Armed School Officials and Fatal and Nonfatal Gunshot Injuries During Mass School Shootings, United States, 1980-2019. JAMA Network Open, 4(2), e2037394.#x2013;e2037394. PubMed. https://doi.org/10.1001/jamanetworkopen.2020.37394

Peterson, J., Erickson, G., Knapp, K., & Densley, J. (2021). Communication of Intent to Do Harm Preceding Mass Public Shootings in the United States, 1966 to 2019. JAMA Network Open, 4(11), e2133073.#x2013; e2133073. PubMed. https://doi.org/10.1001/jamanetworkopen.2021.33073

Pilkington, E. (2022, May 25). US reels after massacre in fourth-grade classroom leaves 21 dead. The Guardian. https://www.theguardian.com/us-news/2022/may/25/biden-reaction-uvalde-school-shooting

Post, L., Mason, M., Singh, L. N., Wleklinski, N. P., Moss, C. B., Mohammad, H., Issa, T. Z., Akhetuamhen, A. I., Brandt, C. A., Welch, S. B., & Oehmke, J. F. (2021). Impact of Firearm Surveillance on Gun Control Policy: Regression Discontinuity Analysis. JMIR Public Health and Surveillance, 7(4), e26042. https://doi.org/10.2196/26042

Reeping, P. M., Cerdá, M., Kalesan, B., Wiebe, D. J., Galea, S., & Branas, C. C. (2019). State gun laws, gun ownership, and mass shootings in the US: cross sectional time series. BMJ (Clinical Research Ed.), 364, l542–l542. PubMed. https://doi.org/10.1136/bmj.l542

Reeping, P. M., Klarevas, L. J., Rajan, S., Rowhani-Rahbar, A., Heinze, J., Zeoli, A. M., Goyal, M. K., Zimmerman, M., & Branas, C. C. (2022). State firearm laws, gun ownership, and K-12 school shootings: Implications for school safety. Journal of School Violence, 21(2), 132–146. https://doi.org/10.1080/15388220.2021.2018332

Rees, C. A., Lee, L. K., Fleegler, E. W., & Mannix, R. (2019). Mass School Shootings in the United States: A Novel Root Cause Analysis Using Lay Press Reports. Clinical Pediatrics, 58(13), 1423–1428. https://doi.org/10.1177/0009922819873650

Ruderman, D., & Cohn, E. G. (2021). Predictive Extrinsic Factors in Multiple Victim Shootings. The Journal of Primary Prevention, 42(1), 59–75. PubMed. https://doi.org/10.1007/s10935-020-00602-3

Siegel, M., Goder-Reiser, M., Duwe, G., Rocque, M., Fox, J. A., & Fridel, E. E. (2020). The relation between state gun laws and the incidence and severity of mass public shootings in the United States, 1976-2018. Law and Human Behavior, 44(5), 347–360. https://doi.org/10.1037/lhb0000378

Silva, J. R. (2022). Global mass shootings: Comparing the United States against developed and developing countries. International Journal of Comparative and Applied Criminal Justice, 1–24. https://doi.org/10.1080/01924036.2022.2052126

Silva, J. R., Capellan, J. A., Schmuhl, M. A., & Mills, C. E. (2021). Gender-Based Mass Shootings: An Examination of Attacks Motivated by Grievances Against Women. Violence against Women, 27(12–13), 2163–2186. https://doi.org/10.1177/1077801220981154

Silver, J., & Silva, J. R. (2022). A Sequence Analysis of the Behaviors and Experiences of the Deadliest Public Mass Shooters. Journal of Interpersonal Violence, 8862605221078818. https://doi.org/10.1177/08862605221078818

Williams, A. (2022, June 22). US Senate unveils bipartisan gun control bill. DW. https://www.dw.com/en/us-agreement-on-gun-control/a-62223101

Wintemute, G. J., Pear, V. A., Schleimer, J. P., Pallin, R., Sohl, S., Kravitz-Wirtz, N., & Tomsich, E. A. (2019). Extreme Risk Protection Orders Intended to Prevent Mass Shootings: A Case Series. Annals of Internal Medicine, 171(9), 655–658. https://doi.org/10.7326/M19-2162

